# A Deep Lightweight Convolutional Neural Network for Detecting Artifacts in Continuous EEG Signals

**DOI:** 10.1101/2025.10.28.25338681

**Authors:** Evans Nyanney, Parthasarathy D. Thirumala, Shyam Visweswaran, Zhaohui Geng

## Abstract

**Objective:** This study aimed to develop and validate a system of specialized deep lightweight convolutional neural networks (CNN) to accurately detect specific artifact classes and demonstrate their advantage over traditional rule-based methods.

**Methods:** Three distinct CNN systems were trained on the Temple University Hospital EEG Artifact Corpus to identify eye movement, muscle-related and non-physiological artifacts, with each system optimized for an ideal temporal window size. The performance of the proposed system was compared with standard rule-based clinical detection methods in a held-out test set.

**Results:** The CNN systems significantly outperformed rule-based methods, with F1-score improvements ranging from +11.2% to +44.9%. Importantly, the results revealed distinct optimal temporal window lengths for each artifact type: 20s for eye movements (ROC AUC 0.975%), 5s for muscle activity (Accuracy 93.2%), and 1s for non-physiological artifact(F1-score 77.4%).

**Conclusion:** The results show that specialized, artifact-specific CNNs provide a more consistent and accurate solution for automated EEG artifact detection than traditional rule-based approaches

**Significance:** This study establishes a new benchmark for automated EEG quality control by validating one of the first open-source, specialized CNN systems for three distinct artifact classes, both high sensitivity and specificity.

## 1. Introduction

Continuous scalp electroencephalography (EEG) remains the primary diagnostic tool for detecting seizures, status epilepticus, and cerebral ischemia in critical care settings. In these environments, most events are non-convulsive or subtle, making continuous monitoring essential for identifying cases that are not clinically apparent but require immediate intervention (Sharma, Nunes and Alkhachroum, 2022; Claassen, Mayer, Kowalski, Emerson and Hirsch, 2004; Mumtaz, Rasheed and Irfan, 2021; Prakash and Kumar, 2024). However, the diagnostic accuracy of EEG is limited by signal contamination from both physiological sources (such as eye movements, muscle activity, chewing, and shivering) and non-physiological sources (including electrode pops, static, and lead artifacts) that not only obscure or mimic pathological EEG patterns but also create substantial workload for neurologists, EEG technologists, and critical care physicians (Jiang, Bian and Tian, 2019; Britton, Frey, Hopp et al., 2016; Urigüen and Garcia-Zapirain, 2015; Islam, Rastegarnia and Yang, 2016). The issue of EEG signal contamination is exacerbated by the increased use of rapid application EEG caps and headbands, especially when applied by untrained personnel on patients with scalp abnormalities(Amin, Nascimento, Karakis, Schomer and Benbadis, 2023) Foundational understanding of EEG artifact characteristics was established through seminal works characterizing spectral properties and topographical distributions of various types of contamination (Goncharova, McFarland, Vaughan and Wolpaw, 2003; Whitham, Pope, Fitzgibbon, Lewis, Clark, Loveless, Broberg, Wallace, DeLosAngeles, Lillie, Hardy, Fronsko, Pulbrook and Willoughby, 2007). This research has led to the development of automated detection frameworks like FASTER (Nolan, Whelan and Reilly, 2010). While traditional approaches including blind-source separation, adaptive filtering, and wavelet decompositions provide partial solutions, they have typically been validated only on small laboratory datasets and lack adaptability across diverse patient populations and recording conditions (Islam et al., 2016; Saba-Sadiya, Chantland, Alhanai, Liu and Ghassemi, 2021; Kalita, Deb and Das, 2024). Recent deep learning approaches have shown promise but still face challenges in clinical deployment due to the incorrect “one-size-fits-all” assumption that artifact characteristics are similar across subjects and tasks (Yu, Li, Zhou, Wang and Wang, 2024; Delorme, Sejnowski and Makeig, 2007; Prasad, Chanamallu and Prasad, 2021).

Our objectives in addressing these challenges are as follows: (1) to develop distinct deep convolutional neural network (CNN) models for eye movements, muscle artifacts, and non-physiological artifacts; (2) to evaluate whether these artifact-specific models outperform single-model approaches; (3) to compare CNN-based methods with rule-based clinical standards; and (4) to provide open-source tools and evaluation frameworks for clinical validation. We hypothesize that models tailored to specific artifacts leveraging the unique characteristics of each type will offer improved diagnostic accuracy. Upon completion, we will release our models, evaluation framework, data splits, and software code under open-source license to facilitate integration of robust artifact removal in future EEG studies and real-time monitoring systems.( See Fig. 1).

**Figure 1:**
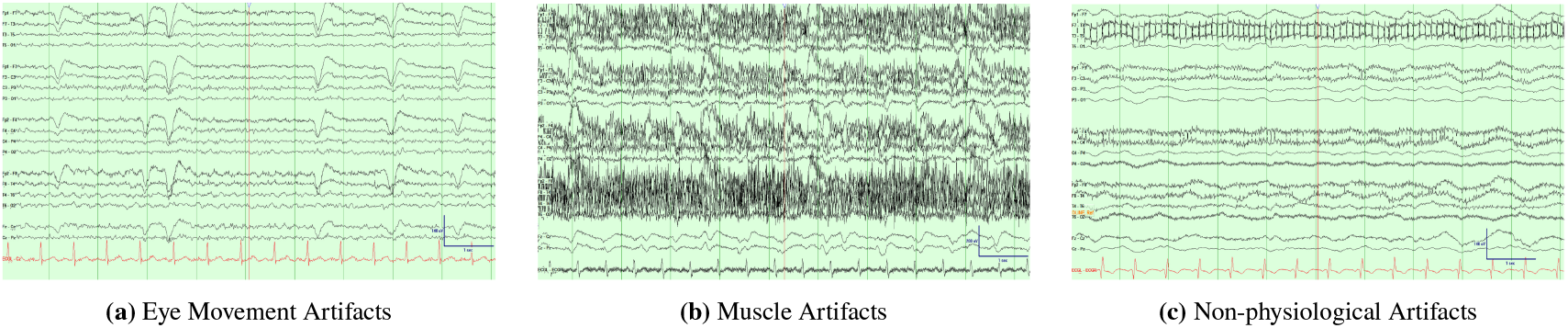
Examples of three main EEG artifact types used in this study. (a) Eye movement artifacts showing smooth, low-frequency deflections with moderate amplitude, (b) muscle artifacts showing high-frequency, irregular activity with higher amplitude, and (c) non-physiological artifacts presenting as sudden, transient spikes characteristic of electrode pops (Amin et al., 2023).

## 2. Materials and Methods

### 2.1. Study Design and Reporting Framework

This study follows a diagnostic accuracy design for developing and validating artifact detection algorithms. We adhere to the TRIPOD+AI (Transparent Reporting of a multivariable prediction model for Individual Prognosis Or Diagnosis + Artificial Intelligence) guidelines for prediction model development (Collins, Moons, Dhiman et al., 2024) and STARD (Standards for Reporting of Diagnostic Accuracy Studies) recommendations for diagnostic test accuracy studies (Bossuyt, Reitsma, Bruns et al., 2015).

### 2.2. Data Description

The development and evaluation datasets used in this study were obtained from the Temple University Hospital (TUH) EEG Corpus(Obeid and Picone, 2016), specifically the edf/01_tcp_ar subset, which represents routine clinical care data collected during standard EEG monitoring procedures from 2002 to 2016. We selected this dataset for several reasons: (1) it features expert-annotated artifact labels with high agreement among the annotators (*κ* > 0.8) (The TUH EEG Corpus, 2020), (2) it offers a diverse range of artifact types and recording conditions, representative of real clinical settings, and (3) it follows standardized collection procedures that ensure data quality and consistency.

The study setting was Temple University Hospital, a tertiary care academic medical center in Philadelphia, Pennsylvania, USA. The dataset comprises 310 curated recordings from patients undergoing routine clinical EEG monitoring across various units, including the epilepsy monitoring unit, the neurointensive care units, and the general neurology units. These recordings were collected during standard clinical workflows, including epilepsy monitoring, sleep studies, and neurological assessments, making the developed models applicable to real-world clinical scenarios.

Participants were included in the study if they: (1) underwent routine clinical EEG monitoring at Temple University Hospital between 2002 and 2016, (2) had recordings of sufficient duration and quality for artifact analysis, and (3) had comprehensive artifact annotations completed by expert neurophysiologists. No specific exclusion criteria were used beyond standard clinical contraindications for EEG monitoring. All recordings were collected during standard clinical care, with patients receiving routine medical treatments as indicated by their clinical conditions. The artifact annotations were completed by expert neurophysiologists who were blinded to the automated detection algorithms.

#### 2.2.1. Artifact Distribution and Characteristics

The data set contains a total of 158,884 artifact annotations across 19 categories(see Table 1), with muscle (30. 4%), eye movement (23. 9%) and electrode artifacts (20.1%) artifacts represent the primary sources of contamination. Both individual and combined artifacts (e.g., eye + muscle, chewing + electrode) provide a realistic representation of complex patterns encountered in clinical practice. Artifact durations range from brief electrode pops (mean 0.165 hours) to prolonged muscle artifacts (mean 0.260 hours), which capture the diverse temporal characteristics important for developing detection models that can effectively handle both transient and continuous artifacts.

**Table 1.** Distribution of EEG artifact types in the TUH dataset with mean duration in hours.

| # | Artifact | Count | % | Mean Duration (hr) | # | Artifact | Count | % | Mean Duration (hr) |
| --- | --- | --- | --- | --- | --- | --- | --- | --- | --- |
| 1 | Muscle | 48,347 | 30.4 | 0.260 | 10 | Generalized seizure | 871 | 0.5 | 0.157 |
| 2 | Eye movement | 38,004 | 23.9 | 0.168 | 11 | Eye + chewing | 864 | 0.5 | 0.111 |
| 3 | Electrode | 31,986 | 20.1 | 0.176 | 12 | Chewing + muscle | 243 | 0.2 | 0.080 |
| 4 | Eye + muscle | 18,455 | 11.6 | 0.172 | 13 | Complex seizure | 198 | 0.1 | 0.239 |
| 5 | Muscle + electrode | 6,949 | 4.4 | 0.201 | 14 | Shivering | 180 | 0.1 | 0.067 |
| 6 | Chewing | 6,482 | 4.1 | 0.121 | 15 | Electrode pop | 172 | 0.1 | 0.165 |
| 7 | Eye + electrode | 2,410 | 1.5 | 0.185 | 16 | Chewing + electrode | 152 | 0.1 | 0.168 |
| 8 | Background noise | 2,374 | 1.5 | 0.118 | 17 | Tonic-clonic seizure | 147 | 0.1 | 0.142 |
| 9 | Focal seizure | 1,004 | 0.6 | 0.131 | 18 | Eye + shivering | 45 | <0.1 | 0.092 |

**Dataset information:** Total annotations: 158,884 across 310 EEG recordings from the edf/01\_tcp\_ar dataset. Durations are the mean artifact start times converted from seconds to hours. Artifacts are classified into physiological (eye movement, muscle, chewing, shivering), technical (electrode, background noise), pathological (seizures), and combined categories.

#### 2.2.2. Data Preprocessing

The raw EEG data from the TUH EEG Corpus required systematic pre-processing to prepare it for training the artifact detection model. We processed 150 EEG recordings that were selected from the 310 available recordings from the edf/01_tcp_ar files. We standardized the variable sampling rates, which ranged from 250 to 1000 Hz, and the channel configurations, which varied from 27 to 36 channels, to uniform specifications. This standardization was done while preserving the original temporal characteristics essential for deep learning models (see Fig. 2).

**Figure 2:**
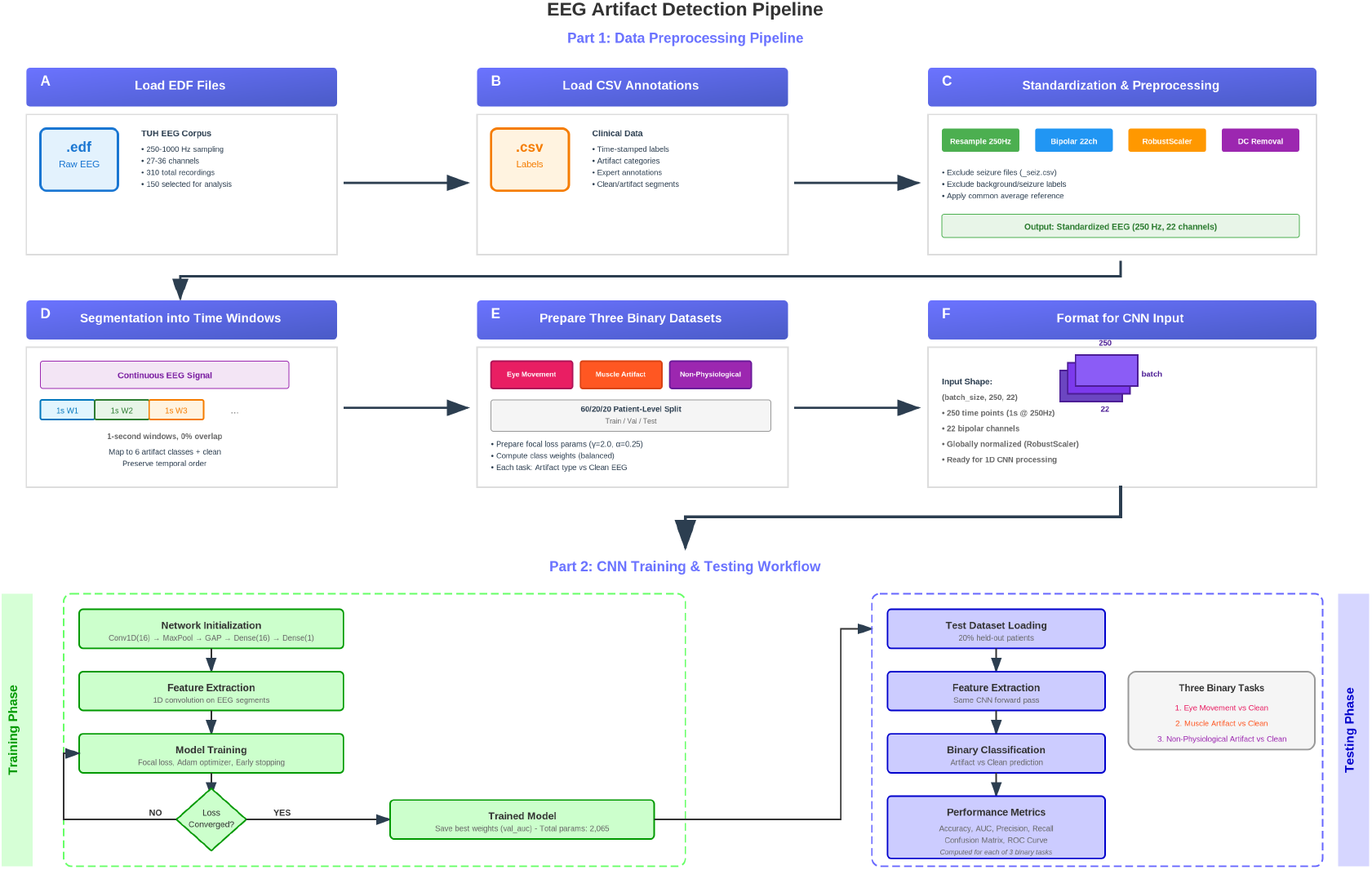
EEG preprocessing pipeline. The process involves: (a) loading EDF files with raw EEG data from 310 recordings (sample of 150), (b) loading CSV annotations with artifact labels, (c) standardization and preprocessing to 250 Hz and 22 channels with seizure file exclusions, (d) segmentation into adaptive non-overlapping windows, (e) preparation of three binary datasets with 60/20/20 patient-level splits and focal loss parameters, and (f) formatting into 3D tensors with global normalization for direct CNN input.

#### 2.2.3. Signal Standardization

All recordings underwent systematic standardization, which included the following steps:(1) resampling to 250 Hz, (2) converting to a standardized 22-channel bipolar montage using canonical electrode pairs (FP1-F7, F7-T3, T3-T5, T5-O1, FP2-F8, F8-T4, T4-T6, T6-O2, A1-T3, T3-C3, C3-CZ, CZ-C4, C4-T4, T4-A2, FP1-F3, F3-C3, C3-P3, P3-O1, FP2-F4, F4-C4, C4-P4, P4-O2), and (3) applying bandpass filtering (1-40 Hz) and notch filtering (50/60 Hz) to remove line noise and artifacts outside the desired frequency range. Although the standard clinical EEG range extends to 70 Hz, the 1-40 Hz range was chosen to specifically target cerebral activity and minimize potential signal contamination by high-frequency muscle (EMG) artifacts, which could otherwise lead to an inflated false positive rate.

#### 2.2.4. Referencing and Normalization

After filtering, we applied average referencing to reduce common-mode noise and removed DC offsets by subtracting the mean value from each channel. We conducted global normalization using RobustScaler across all channels and timepoints, preserving relative amplitude relationships between channels while standardizing the input range for deep learning model training (Lawhern, Solon, Waytowich, Gordon, Hung and Lance, 2018). These preprocessing steps are important to improve the signal-to-noise ratio and to scale the data appropriately for stable model training.

#### 2.2.5. Adaptive Segmentation and Labeling

We segmented each recording into non-overlapping windows of 1, 3, 5, 10, 20, or 30 seconds to create consistent inputs for CNN training. This method maximizes sample independence and preserves the temporal resolution needed for artifact detection. Conducting a structured evaluation across multiple durations is an important step, as the optimal temporal window for detecting an artifact can vary significantly depending on its characteristic timescale.

For each segment, we extracted the corresponding artifact category from the annotation files and labeled the original 19 categories to six artifact classes and a non-artifact class (seven classes): eye movement (EYEM, 0), muscle activity (MUSC, 1), electrode artifact (ELEC, 2), chewing (CHEW, 3), shivering (SHIV, 4), non-physiological artifact (ELPP, 5), and non-artifact segments (6). This adaptive approach captures both brief artifact events (including sub-second events within a 1-second window) and longer patterns, which ensure appropriate temporal resolution for each type of artifact.

#### 2.2.6. Dataset Preparation

We prepared three datasets with binary classes for the detection of specific artifacts as follows. Segments consisting seizure and background categories were excluded during preprocessing.

##### Eye movements

This dataset included segments of class eye movement (0) that were labeled positive and segments of class non-artifact (6) that were labeled negative, focusing on transient, low-frequency frontal activity (blinks, saccades) distinct from EMG and non-physiological noise.

##### Muscle-related artifacts

This dataset included seg-ments of class muscle artifact (1), chewing (3), and shivering (4) that were labeled positive and segments of class non-artifact (6) that were labeled negative, due to shared broad-band, high-frequency content and similar impact on signal readability.

##### Non-physiological artifacts

This dataset included segments of class electrode artifact (2) and non-physiological artifact (5) that were labeled positive and segments of class non-artifact (6) that were labeled negative, to capture step changes, flatlines, impedance pops, and line-related contamination.

Each dataset used the standardized 22-channel bipolar montage with non-overlapping segments at 250 Hz. The dataset included 105 patients with 150 recordings, split into training (63 patients, 60%), validation (21 patients, 20%), and test (21 patients, 20%) sets using patient-level splitting to prevent data leakage. Class imbalance was handled using focal loss (α = 0.25, γ = 2.0), a technique that gives more weight to hard-to-classify segments (Lin, Goyal, Girshick, He and Dollár, 2017). To ensure an unbiased evaluation, thresholds were selected on the validation set to maximize Youden’s J statistic and then applied unchanged to the test set(Powers, 2011).

### 2.3. Deep Learning Method

The detection of artifacts in EEG signals presents significant challenges due to complex temporal patterns and the high-dimensionality of the data. Traditional rule-based methods, which rely on predefined thresholds and frequency-domain features, often struggle to capture the complex temporal dependencies associated with EEG artifacts (Delorme et al., 2007). However, increasing model complexity does not necessarily lead to improved accuracy; it can even cause performance decline without solving the high variance-high bias problem. To address these issues, we developed a deep lightweight CNN architecture for EEG artifact detection that achieves better performance while using fewer trainable parameters. The architecture employs multiple convolutional layers that progressively extract hierarchical features from raw EEG signals, while using depthwise separable convolutions, reduced filter sizes, and efficient channel configurations to minimize computational complexity and memory requirements.

Our framework uses a single CNN designed for one-dimensional temporal EEG data. It incorporates convolutional layers, max-pooling layers, and global average pooling layers to extract features from the raw temporal information and classify them into various artifact categories.. All layer parameters are optimized together by minimizing classification error across the training set.

The convolutional layer operates across the temporal dimension of the input EEG segments, generating activation maps that capture temporal features relevant to artifact detection. The weights*ω*_*ij*_ represent the filters connecting the input channels (*i*) and the output feature maps (*j*), while *β*_*j*_ represents bias. The convolution operation is:

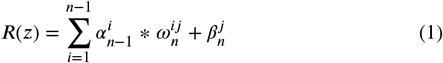

We used ReLU activation for its non-linearity, computational speed, and ability to handle non-negative inputs:

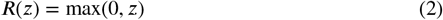

Max-pooling reduces the temporal size of feature maps, which helps decrease the number of parameters and computational costs while preventing overfitting. Global average pooling computes the average value of each feature map across the temporal dimension, creating a single feature vector and serving as a regularizer (Lin, Chen and Yan, 2013). The framework was optimized through an iterative process balancing computational complexity and performance, using patient-level splitting with cross-validation to evaluate and select the best configuration.

#### 2.3.1. Loss Function and Optimization

We used the Adam optimizer with focal loss to address severe class imbalance inherent in EEG artifact detection by assigning higher weights to hard-to-classify examples:

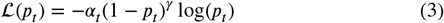

where *p*_*t*_ is the predicted probability for the true class, *α*_*t*_ is the weighting factor for class *t*, and *γ* is the focusing parameter. We use *α* = 0.25 and *γ* = 2.0 to effectively handle class imbalance while maintaining focus on hard-to-classify examples.

#### 2.3.2. Model Architecture and Training

Inputs were multichannel EEG segments in a standardized 22-channel bipolar montage. The temporal length *T* was adaptive and inferred at runtime, which was the same design to operate across different segment lengths without structural changes. The deployed model used a lightweight one-dimensional architecture: Conv1D layer (16 filters, kernel size 5, ReLU, same padding), MaxPooling1D (pool size 2), global average pooling over the temporal dimension, and a two-layer classifier (Dense 16, ReLU; Dense 1, sigmoid). This lightweight design without BatchNormalization or Dropout was chosen based on empirical performance comparisons(Lawhern et al., 2018).

#### 2.3.3. Training

Supervised training used binary targets per classifier with mini-batch optimization (batch size 128, maximum 200 epochs) and shuffled batches. Learning-rate scheduling was based on validation performance, retaining the best weights by validation loss. Decision thresholds were selected on the validation set to maximize Youden’s *J* statistic, fixed specificity, or maximize the true positive rate (TPR) with the false positive rate FPR ≤ 0.10, then applied unchanged to the test set(Powers, 2011). Model fitting used GPU acceleration when available, with CPU preprocessing. Random seeds were fixed for numerical reproducibility, with training proceeding on CPU when a GPU was unavailable, using unchanged hyperparameters.

### 2.4. Performance Analyses

We compared the performance of the CNN with rule-based artifact detection methods across three binary classification tasks: eye movement, muscle artifact, and nonphysiological artifact detection. We computed standard classification metrics (accuracy, precision, recall, F1-score, sensitivity, specificity) and specialized metrics for imbalanced datasets, with F1-score, which serves as the primary optimization criterion during threshold selection (Saito and Rehmsmeier, 2015).

For performance evaluation, we utilized two key metrics: AUC ROC (Area Under the Receiver Operating Characteristic Curve), which measures the model’s ability to distinguish between artifact and clean segments across all classification thresholds, with values ranging from 0 to 1 where 1.0 indicates perfect classification, and pROC@0.1 (Partial ROC at 0.1 False Positive Rate), which evaluates performance specifically at low false positive rates by measuring the area under the ROC curve up to a false positive rate of 0.1, which gives insight into the model’s performance under strict false positive constraints.

For each classifier, we implemented dual-threshold optimization on the validation set: (1) F1-optimal thresholds determined by maximizing the harmonic mean of precision and recall from precision-recall curves, and three alternative strategies: (2) thresholds maximizing Youden’s J statistic (sensitivity + specificity - 1), (3) thresholds that obtained 95% specificity to prioritize FPRs, and (4) thresholds that maximized the FPR ≤ 0.1 constraint. All optimizations were performed on the validation sets to prevent test data overfitting.

#### 2.4.1. Prevalence-Adjusted Metrics

To address the imbalance of the data set, we computed the prevalence adjusted area under the precision recall curve (PR AUC) by adjusting the precision values to a target prevalence of 5%:

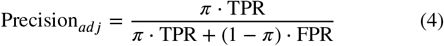

where *π* = 0.05 represents target prevalence, TPR is true positive rate, and FPR is false positive rate. Additionally, we calculated partial ROC-AUC for FPR ≤ 0.1, normalized by the FPR limit to provide clinically performance measures focused on low FPRs (He and Garcia, 2009).

##### 2.4.2. Comparison with Standard Methods

Performance comparisons between CNN and rule-based methods were conducted using all evaluation metrics. Performance differences were evaluated using a threshold-based approach, with differences > 0.01 considered meaningful for distinguishing between methods, while differences < 0.01 were considered ties, indicating equivalent performance. Analysis was performed separately for each artifact detection task.

## 3. Results

### 3.1. Segment-level Artifact Detection

#### Eye Movement Detection

CNN-based eye movement detection performance in different segment sizes achieved ROC AUC values ranging from 0.959 to 0.975, with peak performance in 20-second segments(see Table 2). The 5-second segment provided an optimal balance between accuracy (92.5%) and specificity (94.0%), while the 20-second segments achieved the highest ROC AUC (0.975) and F1 score (0.905). The 30-second segment demonstrated superior precision (95.1%) and pROC@0.1 (0.088), which indicates excellent performance at low false positive rates. The result indicates that eye movement artifacts benefit from a longer temporal context for reliable detection.

**Table 2.** Eye Movement - Window Optimization.

| Artifact Type | Window Length | Performance Metrics |  |  |  |  |  |  |
| --- | --- | --- | --- | --- | --- | --- | --- | --- |
|  |  | ROC AUC | pROC@0.1 | Acc | Prec | Rec | Spec | F1 |
| Eye Movement | 1s | 0.959 | 0.0767 | 89.8 | 73.5 | 89.8 | 89.8 | 80.8 |
|  | 3s | 0.972 | 0.0832 | 91.1 | 79.0 | **93.0** | 90.4 | 85.4 |
|  | 5s | 0.968 | 0.0825 | **92.5** | 86.9 | 89.1 | **94.0** | 88.0 |
|  | 10s | 0.971 | 0.0827 | 90.9 | 86.4 | 89.3 | 91.8 | 87.9 |
|  | 20s | **0.975** | 0.0854 | 91.6 | 94.9 | 86.5 | 96.0 | **90.5** |
|  | 30s | 0.967 | **0.0879** | 89.3 | **95.1** | 83.2 | 95.6 | 88.8 |

**Table description:** Held-out test results at Youden's $J$ operating point (chosen on validation; applied unchanged to test). Metrics: **Acc** = accuracy, **Prec** = precision, **Rec** = recall (sensitivity), **Spec** = specificity, **F1** = F1-score, **ROC AUC** = area under ROC (threshold-independent), **pROC@0.1** = ROC AUC for FPR $\leq 0.10$ (normalized). Windows are non-overlapping; sample counts at 250 Hz: 1 s = 250, 3 s = 750, 5 s = 1,250, 10 s = 2,500, 20 s = 5,000, 30 s = 7,500. Best per column is highlighted; ties not marked.

#### Muscle Artifact Detection

CNN-based muscle artifact detection results varied significantly with segment length, with ROC AUC values ranging from 0.931 to 0.977 (see Table 3). Intermediate segment lengths (3-10 seconds) outperformed both shorter and longer segments, with 3-second segments achieving the highest ROC AUC (0.977) and 5-second segments providing peak accuracy (93.2%). The 10-second segment shows superior F1-score (0.863) and pROC@0.1 (0.089) performance. Unlike eye movements, muscle artifacts showed optimal detection in moderate temporal context, with performance declining at very short (1 second) and long (20-30 second) durations.

**Table 3.**
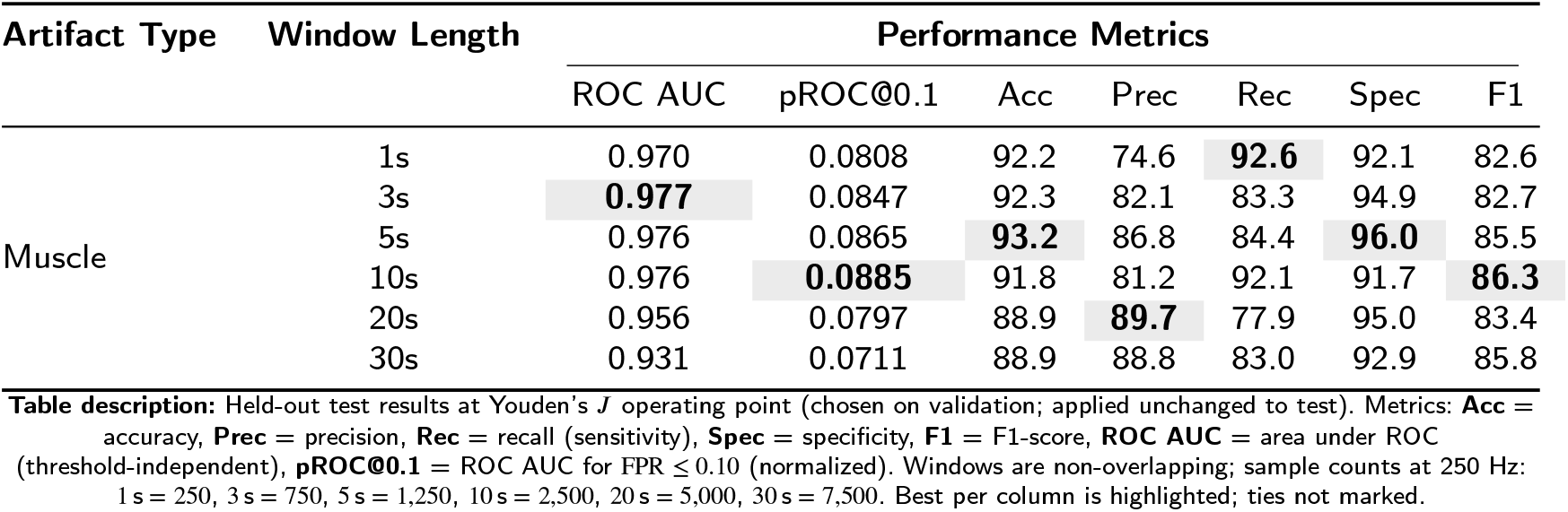
Muscle Artifact - Window Optimization.

#### Non-Physiological Artifact Detection

CNN-based detection of non-physiological artifacts was highly dependent on the size of the segment, with performance varying significantly between the durations tested (see Table 4). A clear pattern emerged in which shorter segments (1-3 seconds) consistently outperformed longer ones and performance declined as the segment size increased. Peak performance was achieved with 1-second segments, producing the best results for precision, specificity, F1 score, and pROC@0.1. This confirms that the transient nature of non-physiological events require shorter temporal segments for optimal detection.

**Table 4.** Non-Physiological - Window Optimization.

| Artifact Type | Window Length | Performance Metrics |  |  |  |  |  |  |
| --- | --- | --- | --- | --- | --- | --- | --- | --- |
|  |  | ROC AUC | pROC@0.1 | Acc | Prec | Rec | Spec | F1 |
| Non-Physiological | 1s | **0.950** | **0.0775** | **96.1** | 80.3 | 74.6 | **98.2** | **77.4** |
|  | 3s | 0.940 | 0.0735 | 93.7 | 69.2 | **80.0** | 95.5 | 74.2 |
|  | 5s | 0.930 | 0.0756 | 92.7 | 69.9 | 78.2 | 94.9 | 73.8 |
|  | 10s | 0.888 | 0.0645 | 91.5 | 77.8 | 71.8 | 95.7 | 74.7 |
|  | 20s | 0.886 | 0.0642 | 88.8 | **87.4** | 64.8 | 96.9 | 74.5 |
|  | 30s | 0.865 | 0.0582 | 84.9 | 82.5 | 64.8 | 93.8 | 72.6 |

**Table description:** Held-out test results at Youden's $J$ operating point (chosen on validation; applied unchanged to test). Metrics: **Acc** = accuracy, **Prec** = precision, **Rec** = recall (sensitivity), **Spec** = specificity, **F1** = F1-score, **ROC AUC** = area under ROC (threshold-independent), **pROC@0.1** = ROC AUC for FPR $\leq 0.10$ (normalized). Windows are non-overlapping; sample counts at 250 Hz: 1 s = 250, 3 s = 750, 5 s = 1,250, 10 s = 2,500, 20 s = 5,000, 30 s = 7,500. Best per column is highlighted; ties not marked.

### 3.2. Effect of Window Size on Detection

We conducted an optimization of the segment size to determine the ideal temporal resolution for each artifact classifier. Our main focus was on specificity (Goncharova et al., 2003; Whitham et al., 2007; Nolan et al., 2010), while we also considered the F1-score, ROC AUC, and accuracy as secondary metrics.

The eye movement detector achieved optimal performance with 20-second segments (ROC AUC: 0.975, F1-score: 90.5%) (see Table 2).

The Muscle artifact detector performed best with 5-second segments (accuracy: 93.2%, specificity: 96.0%) (see Table 3).

The Non-physiological artifact detector achieved optimal performance with 1-second segments (ROC AUC: 0.950, accuracy: 96.1%, specificity: 98.2%, F1-score: 77.4%) (see Table 4).

The results highlight specific temporal requirements for different types of artifacts: non-physiological artifacts are transient and are most effectively detected with a 1-second context, muscle artifacts require a moderate context of 5 seconds, while eye movements benefit from a longer temporal context of 20 seconds. Using Youden’s index for optimal threshold selection, our final models show strong performance across all artifact types (see Table 5). The eye movement detector achieved 94.2% sensitivity and 89.3% specificity, the muscle artifact detector reached 89.5% sensitivity and 95.1% specificity, while the non-physiological detector attained 87.8% sensitivity and 88.9% specificity. The Figure 3 shows the confusion matrices for each detector at their optimal configurations, which illustrates the classification performance on the test set.

**Table 5.** Final Model Performance Summary at Optimal Window Sizes.

| Artifact Detector | Optimal Window | Performance Metrics |  |  |  |  |  |  |  |
| --- | --- | --- | --- | --- | --- | --- | --- | --- | --- |
|  |  | ROC AUC | pROC@0.1 | Acc (%) | Prec (%) | Rec (%) | Spec (%) | F1 (%) | Threshold |
| Eye Movement | 20s | 0.975 | 0.0854 | 91.6 | 94.9 | 86.5 | 96.0 | 90.5 | 0.355 |
| Muscle Artifact | 5s | 0.976 | 0.0865 | 93.2 | 86.8 | 84.4 | 96.0 | 85.5 | 0.418 |
| Non-Physiological | 1s | 0.950 | 0.0775 | 96.1 | 80.3 | 74.6 | 98.2 | 77.4 | 0.359 |

**Figure 3:**
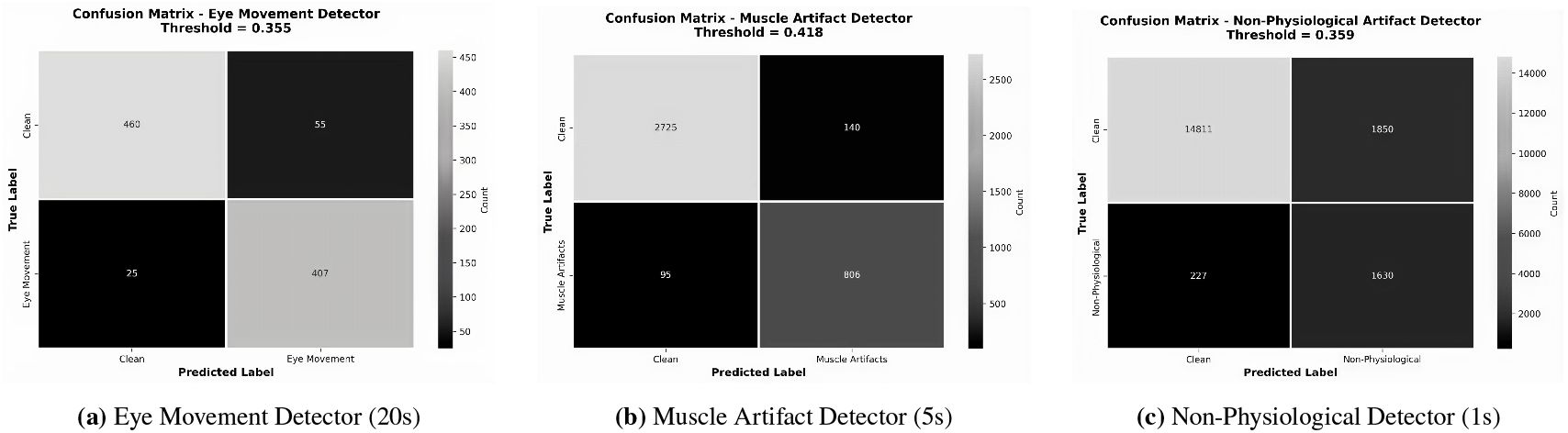
Confusion matrices for artifact detectors using optimal window sizes with Youden’s index thresholds.

### 3.3. Comparative Analysis

Comparison of performance between CNN and rule-based methods in all types of artifacts and window sizes (see Table 6). For eye movement detector, the CNN showed improvements in the F1 score from +11. 2% (10-second windows) to +15. 9% (1-second windows). The CNN had higher accuracy, precision, recall, and F1-score across all window sizes, with rule-based methods only better in specificity at 10-second windows.

**Table 6.** Performance metrics (%) for Deep Lightweight CNN vs Rule-Based artifact detection across six temporal windows. Best values per metric in **bold** with color coding.

| Artifact | Window | Deep Lightweight CNN | | | | | Rule-Based Method | | | | | $\Delta F1$ |
| --- | --- | --- | --- | --- | --- | --- | --- | --- | --- | --- | --- | --- |
|  |  | Acc | Prec | Rec | F1 | Spec | Acc | Prec | Rec | F1 | Spec |  |
| Eye Movement | 1s (n=30,699) | **89.8** | **73.5** | **89.8** | **80.8** | **89.8** | 82.1 | 61.4 | 68.9 | 64.9 | 86.3 | +15.9 |
|  | 3s (n=9,996) | **91.1** | **79.0** | **93.0** | **85.4** | **90.4** | 84.2 | 71.2 | 73.1 | 72.1 | 88.5 | +13.3 |
|  | 5s (n=5,868) | **92.5** | **86.9** | **89.1** | **88.0** | **94.0** | 84.2 | 75.8 | 72.0 | 73.9 | 89.7 | +14.1 |
|  | 10s (n=2,811) | **90.9** | **86.4** | **89.3** | **87.9** | 91.8 | 84.0 | 83.6 | 70.7 | 76.6 | **91.9** | +11.2 |
|  | 20s (n=1,303) | **91.6** | **94.9** | **86.5** | **90.5** | **96.0** | 80.9 | 89.6 | 66.1 | 76.0 | 93.5 | +14.4 |
|  | 30s (n=823) | **89.3** | **95.1** | **83.2** | **88.8** | **95.6** | 80.0 | 92.9 | 65.5 | 76.8 | 94.8 | +12.0 |
| Mean |  | 90.9 | 84.2 | 88.5 | 85.2 | 91.9 | 82.6 | 76.4 | 69.3 | 71.6 | 90.0 | +13.5 |
| Muscle/EMG | 1s (n=29,177) | **92.2** | **74.6** | **92.6** | **82.6** | 92.1 | 85.6 | 71.3 | 46.9 | 56.6 | **95.3** | +26.0 |
|  | 3s (n=9,232) | **92.3** | **82.1** | **83.3** | **82.7** | 94.9 | 85.5 | 79.9 | 45.7 | 58.1 | **96.8** | +24.5 |
|  | 5s (n=5,326) | **93.2** | **86.8** | **84.4** | **85.5** | 96.0 | 84.6 | 82.5 | 44.9 | 58.2 | **97.0** | +27.4 |
|  | 10s (n=2,457) | **91.8** | 81.2 | **92.1** | **86.3** | 91.7 | 82.4 | **89.0** | 42.4 | 57.4 | **98.0** | +28.9 |
|  | 20s (n=1,098) | **88.9** | 89.7 | **77.9** | **83.4** | 95.0 | 76.1 | **93.3** | 35.6 | 51.6 | **98.6** | +31.8 |
|  | 30s (n=683) | **88.9** | 88.8 | **83.0** | **85.8** | 92.9 | 73.1 | **97.0** | 34.7 | 51.1 | **99.3** | +34.8 |
| Mean |  | 91.2 | 83.9 | 85.6 | 84.4 | 93.8 | 81.2 | 85.5 | 41.7 | 55.5 | 97.5 | +28.9 |
| Non-Physiological | 1s (n=25,644) | **96.1** | **80.3** | **74.6** | **77.4** | **98.2** | 75.9 | 21.7 | 64.4 | 32.5 | 77.1 | +44.9 |
|  | 3s (n=8,117) | **93.7** | **69.2** | **80.0** | **74.2** | **95.5** | 71.9 | 20.6 | 52.4 | 29.6 | 74.4 | +44.6 |
|  | 5s (n=4,671) | **92.7** | **69.9** | **78.2** | **73.8** | **94.9** | 72.6 | 24.6 | 51.8 | 33.4 | 75.8 | +40.4 |
|  | 10s (n=2,146) | **91.5** | **77.8** | **71.8** | **74.7** | **95.7** | 73.4 | 33.7 | 53.7 | 41.4 | 77.6 | +33.3 |
|  | 20s (n=941) | **88.8** | **87.4** | **64.8** | **74.5** | **96.9** | 71.0 | 42.9 | 47.0 | 44.9 | 79.0 | +29.6 |
|  | 30s (n=588) | **84.9** | **82.5** | **64.8** | **72.6** | **93.8** | 70.1 | 51.8 | 47.3 | 49.4 | 80.3 | +23.2 |
| Mean |  | 91.3 | 77.9 | 72.4 | 74.5 | 95.8 | 72.5 | 32.6 | 52.8 | 38.5 | 77.4 | +36.0 |
| **Overall Performance** |  |  |  |  |  |  |  |  |  |  |  |  |
| DL-CNN superior / Total metrics |  |  | 82 / 90 (91.1%) |  |  |  | Rule-based superior: 8 / 90 (8.9%) |  |  |  |  |  |

**Notes:** (1) Sample sizes (n) shown for each window. Total test samples vary by artifact type due to class imbalance. (2) Acc = Accuracy, Prec = Precision, Rec = Recall, F1 = F1-Score, Spec = Specificity, $\Delta F1$ = DL-CNN F1 - Rule F1. (3) Threshold optimization using Youden's J statistic. (4) Window sizes: 1s = 250 samples, 3s = 750 samples, 5s = 1,250 samples, 10s = 2,500 samples, 20s = 5,000 samples, 30s = 7,500 samples at 250 Hz. All windows are non-overlapping segments extracted from continuous EEG recordings. (5) The Deep Lightweight CNN: 4-layer convolutional network (16-32-64-128 filters) trained without regularization (no dropout, no weight decay) using focal loss ( $\gamma=2$ , $\alpha=0.25$ ) to address class imbalance naturally without artificial class weighting. (6) Rule-based methods: EMG detection via 20-50 Hz power spectral analysis (Goncharova et al., 2003; Whitham et al., 2007); Non-physiological via FASTER variance thresholds (Nolan et al., 2010); Eye movement along frontal low-frequency dominance analysis.

Muscle artifact detection showed the most significant performance differences between the methods. The CNN had substantial improvements in the F1 score from +24. 5% to +34. 8%, with substantial improvements in 30-second windows. Although rule-based methods had higher specificity across all window sizes, the CNN showed superior performance in accuracy, precision, recall, and F1-score, with the performance gap larger at increased window sizes.

Non-physiological artifact detection showed the largest overall performance differences, with F1 score improvements of +23. 2% (30-second windows) to +44.9% (1 1-second windows). The CNN demonstrated superior performance across all metrics, with the performance advantage smaller at longer window sizes but still substantial throughout. These consistent improvements suggest that deep learning approaches better capture the complex temporal patterns present in EEG artifacts.

## 4. Discussion

We compare our findings to established literature on EEG artifact detection. Goncharova et al. identified muscle artifacts by power spectral analysis in the 20-50 Hz range but reported difficulties with specificity in clinical settings (Goncharova et al., 2003). Whitham et al. investigated the frequency properties of muscle artifacts using spectral-based methods and achieved moderate performance in artifact detection (Whitham et al., 2007). Nolan et al. created the FASTER framework, which uses variance thresholds for automated artifact rejection. This framework reflects current clinical standards, boasting an accuracy rate of 85-90%, although it has limited data on specificity (Nolan et al., 2010). Compared to these rule-based methods, our system performs better in terms of overall performance. We report a high specificity of 96.0%-98.2% while achieving a competitive sensitivity of 74.6%-86.5% across artifact types, making it appropriate for real-world applications. Rule-based methods in our evaluation achieved high specificity for muscle artifacts (95.3%-99.3%) but showed poor sensitivity (34.7%-46.9%) and substantially lower F1-scores. Our approach shows F1-score improvements of +11.2% to +44.9% in all types of artifacts. Traditional spectral and variance-based methods showed particularly poor performance for non-physiological artifacts, which is problematic as it leads to significant loss of valuable EEG information. The length of the right window matters: 20s (eye), 5s (muscle), 1s (non-phys). A single general model performs worse than per-artifact models. For clinical use, our models can provide real-time predictions using sliding window approaches. Even though models are trained on longer windows (like 20 seconds), they can still give predictions every second by looking back at the required window length. This addresses clinical needs for frequent artifact detection while keeping the performance benefits of longer temporal contexts during training.

A key contribution is that we are one of the first to develop a CNN model with high sensitivity and specificity for each of the three artifact classes. A limitation of this work is that it is based on a single dataset, and the model needs to be evaluated in other datasets to ensure generalizability.

## 5. Conclusion

We developed a deep lightweight CNN system for automated detection of three artifact classes: eye movement, muscle activity, and non-physiological artifacts. The system consists of three binary detectors that use different time windows and focal loss to handle class imbalance. We evaluated the system through window size tests and compared it with rule-based methods. The CNN system showed better performance than rule-based methods in all types of artifacts. The high specificity achieved by the CNN system is particularly important for clinical applications, as it minimizes false alarms and reduces unnecessary interruptions in patient monitoring. By making our artifact-specific models and evaluation framework publicly available, we provide a valuable resource for the neurophysiology research community. In future work, artifact detection can be used with other EEG detectors, such as seizure or slowing detectors, to improve clinical monitoring systems.

## Data Availability

The data analyzed are from the Temple University Hospital (TUH) EEG Corpus, specifically the edf/01_tcp_ar subset (routine clinical EEG, 2002-2016). Access is available to qualified researchers upon registration and acceptance of the dataset's data-use terms; we are not authorized to redistribute raw TUH data.

https://isip.piconepress.com/projects/nedc/html/tuh_eeg/

## CRediT authorship contribution statement

**Evans Nyanney:** Conceptualization, Methodology, Software, Writing - Original Draft. **Parthasarathy D. Thirumala:** Supervision, Validation, Writing - Review & Editing. **Shyam Visweswaran:** Supervision. **Zhaohui Geng:** Methodology, Formal analysis.

## Notes

### Competing Interest Statement

The authors have declared no competing interest.

### Funding Statement

This study did not receive external funding.

### Author Declarations

Institutional Review Board of the University of Pittsburgh, Human Research Protection Office, waived ethical approval for this work (secondary analysis of de-identified EEG data). Temple University Institutional Review Board approved the original collection of the Temple University Hospital (TUH) EEG Corpus; this study used the edf/01_tcp_ar subset (routine clinical EEG collected 2002-2016).

